# High neutralizing activity against Omicron BA.2 can be induced by COVID-19 mRNA booster vaccination

**DOI:** 10.1101/2022.04.19.22273940

**Authors:** Lidya Handayani Tjan, Koichi Furukawa, Yukiya Kurahashi, Silvia Sutandhio, Mitsuhiro Nishimura, Jun Arii, Yasuko Mori

## Abstract

The VOC of SARS-CoV-2, Omicron (BA.1, BA.1.1, BA.2, or BA.3), is associated with an increased risk of reinfection. BA.2 has become the next dominant variant worldwide. Although BA.2 infection has been shown to be mild illness, its high transmissibility will result in high numbers of cases. In response to the surge of Omicron BA.1 cases, booster vaccination was initiated in many countries. But there is limited evidence regarding the effectiveness of a booster vaccination against BA.2. We collected blood samples from 84 physicians at Kobe University Hospital, Japan, in January 2022 ∼7 months after they had received two BNT162b2 vaccinations and ∼2 weeks after their booster vaccination. We performed a serum neutralizing assay against BA.2 using authentic virus.

Although most of the participants had no or a very low titer of neutralizing antibody against BA.2 at 7 months after two BNT162b2 vaccinations, the titer increased significantly at 2 weeks after the booster vaccination.

## Introduction

The most recently designated VOC of SARS-CoV-2, Omicron, is associated with an increased risk of reinfection^1^. Among four sub-lineages of Omicron (BA.1, BA.1.1, BA.2, and BA.3), BA.1 has spread to >151 countries and is responsible for the greatly increased number of COVID-19 cases worldwide. However, Omicron BA.2 has also been detected in ≥85 countries and became the dominant lineage in 18 countries by mid-February 2022^2^. BA.2 has become the next dominant variant worldwide.

A recent epidemiological study in South Africa suggested that the clinical profile of illness caused by BA.1 infection is similar to that caused by BA.2^3^. Although BA.2 infection has been shown to be mild illness, its high transmissibility will result in high numbers of cases with considerable societal impacts, e.g., greater work absences, including healthcare and public employees.

In response to the surge of Omicron BA.1 cases, booster vaccination was initiated in many countries. We and others have previously reported that BA.1 escapes two doses of BNT162b2 mRNA vaccine-induced neutralization and that a booster (3rd) vaccination is required to induce the neutralizing antibody against BA.1^4,5^. But there is limited evidence regarding the effectiveness of a booster vaccination against BA.2 ^6^.

## Methods

We collected blood samples from 84 physicians at Kobe University Hospital, Japan, in January 2022 (median age 44 years, IQR 33–58) ∼7 months after they had received two BNT162b2 vaccinations and ∼2 weeks after their booster vaccination. We performed a serum neutralizing assay against BA.2 using authentic virus as described^4^. No participants had a history of SARS-CoV-2 infection. The study was approved by the ethical committee of the Kobe University Graduate School of Medicine (approval code: B200200). All participants were recruited with their written consent.

## Results

The results demonstrated that similar to BA.1^4^, most of the participants had no or a very low titer of neutralizing antibody against BA.2 at 7 months after two BNT162b2 vaccinations (GMT 1□18, 95%CI: 1□09–1□27). However, the titer increased significantly at 2 weeks after the booster vaccination (GMT 36□44, 95%CI: 30□53–43□50), p<0□001 (Fig. 1).

**Figure. 1.**
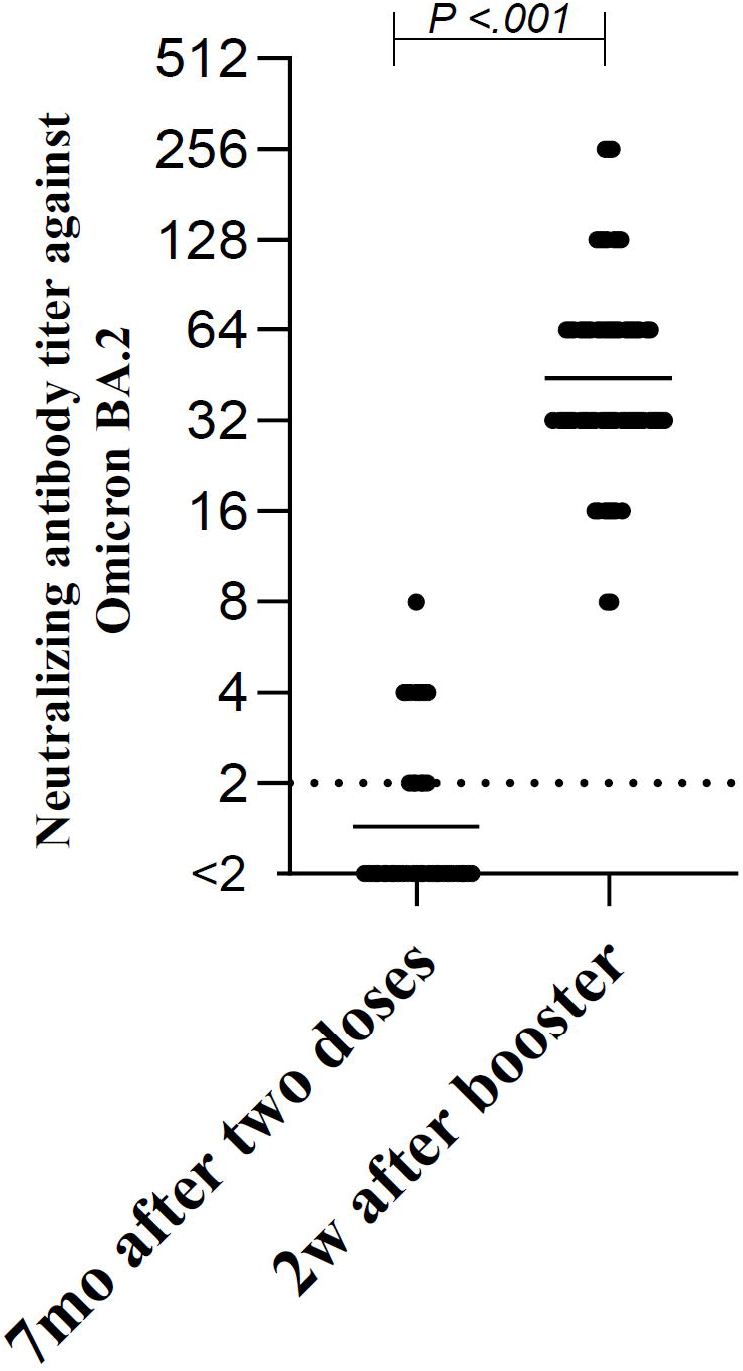
Neutralizing antibody titers against Omicron BA.2 in BNT162b2 mRNA-vaccinated adult males (n=84) at 7 months after they had received two vaccine doses and at 2 weeks after a booster vaccination. The limit of detection is displayed with a dotted horizontal line and the horizontal line shows the geometric mean titer. The titers were compared by the two-sample Wilcoxon rank-sum (Mann-Whitney) test; two-tailed p-values were calculated.

## Discussion

These results indicate that the booster vaccination could induce neutralizing immunity against Omicron BA.2 (as it has against BA.1), and that a booster dose of BNT162b2 mRNA vaccine induces a high cross-neutralizing response against SARS-CoV-2 variants^4^, indicating the booster vaccination is meaningful and hopeful for the suppression of BA.2 pandemic and can activate memory B cells that could produce neutralizing antibodies recognizing epitopes conserved among SARS-CoV-2 variants.

## Data Availability

All data produced in the present work are contained in the manuscript.

## Acknowledgments

We gratefully acknowledge Kazuro Sugimura MD, PhD (Superintendent, Hyogo Prefectural Hospital Agency and Professor, Kobe University) for giving his full support to this study. We also thank the National Institute of Infectious Disease Japan for providing the SARS-CoV-2 Omicron variants BA.2.

## References

1. Altarawneh HN, Chemaitelly H, Hasan MR, et al. Protection against the Omicron Variant from Previous SARS-CoV-2 Infection. N Engl J Med. Feb 9 2022;doi:10.1056/NEJMc2200133

2. World Health Organization (WHO). COVID-19 Weekly Epidemiological Update Edition 80. Updated 22 February 2022. Accessed Mar 15, 2022. https://apps.who.int/iris/bitstream/handle/10665/352199/CoV-weekly-sitrep22Feb22-eng.pdf

3. Wolter N, Jassat W, Walaza S, et al. Early assessment of the clinical severity of the SARS-CoV-2 omicron variant in South Africa: a data linkage study. Lancet. Jan 29 2022;399(10323):437–446. doi:10.1016/S0140-6736(22)00017-4

4. Furukawa K, Tjan LH, Kurahashi Y, et al. Acquired neutralizing breadth against SARS-CoV-2 variants including Omicron after three doses of mRNA COVID-19 vaccination and the vaccine efficacy. medRxiv [preprint]. 2022;doi:10.1101/2022.01.25.22269735 doi:https://doi.org/10.1101/2022.01.25.22269735

5. Nemet I, Kliker L, Lustig Y, et al. Third BNT162b2 Vaccination Neutralization of SARS-CoV-2 Omicron Infection. N Engl J Med. Feb 3 2022;386(5):492–494. doi:10.1056/NEJMc2119358

6. Chen LL, Chu AW, Zhang RR, Hung IF, To KK. Serum neutralisation of the SARS-CoV-2 omicron sublineage BA.2. The Lancet Microbe. Mar 28 2022;doi:10.1016/S2666-5247(22)00060-X

